# The Impact of the President’s Emergency Program for AIDS Relief (PEPFAR) on Children

**DOI:** 10.1101/2023.09.18.23295712

**Authors:** John Stover, Clare F. Flanagan, Yu Teng

## Abstract

**Background:** The AIDS epidemic has had severe impacts on children causing 7.7 million child deaths and orphaning 15 million children. The impact would have been even greater if the PEPFAR program had not provided drugs and services to avert millions of new infections and AIDS-related deaths. This paper estimates the historical impact of PEPFAR support on children and projects the impact through 2030 if the PEPFAR program were to stop in 2024.

**Methods:** We used the Goals RSM HIV simulation model to project counterfactual scenarios of no PEPFAR support during both the historical period (2000-2022) and the future period (2024-2030). We applied the model to 53 countries receiving PEPFAR support.

**Results:** From 2004-2022 the PEPFAR program averted 2.8 million new child HIV infections, 1.5 million child HIV-related deaths and prevented 8.2 million children from becoming AIDS orphans. A continuation of PEPFAR at its current level through 2030 would avert one million new child HIV infections, 460,000 child HIV-related deaths and 2.8 million AIDS orphans.

**Discussion:** The PEPFAR program has been crucial to the success that has been achieved to date in the global fight against AIDS. That success has created an obligation. Continuation of PEPFAR is critical to the effort to achieve the end of AIDS in the coming years.

## Background

The global HIV pandemic has been one of the most serious health challenges the world has faced. The effects have been particularly severe for children. According to UNAIDS, 7.7 million children aged 0-14 have died of AIDS through 2022, 1.5 million children are living with HIV today and over 15 million children under the age of 18 have lost one or both parents due to AIDS. [1] This situation could have been even worse. New child HIV infections peaked at 540,000 (360,000 – 830,000) per year in 1999 and have since declined to 130,000 (90,000 – 210,000) by 2022. Child HIV-related deaths peaked in 2002 at 360,000 (240,000 – 520,000) and have dropped to 84,000 (56,000 – 770,000) today. Progress since 2000 is due to the scale-up of programs to prevent vertical HIV transmissions at birth (VT), antiretroviral therapy (ART) for both adults and children and primary prevention interventions (such as condoms, voluntary medical male circumcision, and peer outreach) that have reduced incidence among adults. Today 82% of pregnant women living with HIV receive antiretroviral drugs (ARVs) to prevent vertical transmission and about 57% of children living with HIV are on ART. [2]

Programs to prevent, treat and mitigate the effects of HIV have been funded by national governments, out-of-pocket payments by individuals and international donors. About 60% of funding for HIV programs in low- and middle-income countries is supplied by international donors, and 58% of those funds come from the United States President’s Emergency Plan for AIDS Relief (PEPFAR). [3] Within PEPFAR-supported countries, the proportion of PEPFAR within national HIV spending is even higher.

Several previous studies have examined the impact of PEPFAR. Bendavid and colleagues [4] and Kates and colleagues [5] found that PEPFAR significantly reduced AIDS-related mortality. In a 2015 study Heaton et al. [6] found that PEPFAR averted 2.9 million HIV infections and 9 million orphans from 2004-2013. Crown et al. [7] estimated the non-health impact of PEPFAR and found significant benefits to economic growth and school enrollment.

Indeed, PEPFAR’s efforts have been monumental in spurring progress towards the UNAIDS 95-95-95 targets with some believing the end of AIDS is achievable as soon as 2030. [8-9] Despite this incredible impact, however, PEPFAR is now under threat and could be defunded as soon as 2024. [10]

The purpose of the work reported here is to estimate the historical impact of PEPFAR on children and to use simulation modeling to project the importance of PEPFAR continuation to ensure progress towards achieving the global AIDS targets and ending the HIV epidemic.

## Data and Methods

### Current PEPFAR coverage

PEPFAR is currently active in 55 low- and middle-income countries. (The full list of countries is available at https://www.state.gov/pepfar-supported-countries-and-regions.) PEPFAR provides a wide variety of services including primary prevention, testing and treatment, mitigation and health system strengthening. For the purpose of this analysis, we focused on five PEPFAR services:

- Prevention of vertical transmission (PVT), which reduces new child infections
- Pediatric ART, which reduces child HIV-related deaths
- Adult ART, which reduces adult AIDS deaths and AIDS orphans
- Voluntary medical male circumcision (VMMC), which reduces adult prevalence of HIV
- Services for key populations, which reduce adult prevalence of HIV

From 2004 to 2022 PEPFAR provided 22 million person-years with PVT, 276 million person-years of ART, 58 million VMMCs and 66 person-years of prevention services to key populations. Information on the volume of each of these services by year and recipient country is available from the PEPFAR website at https://data.pepfar.gov/additionalData.

### Goals RSM simulation model overview

To assess the impact of these services we used the Goals RSM simulation model. The Goals model is implemented for an individual country by using country-specific data for demographic indicators (base year population, fertility, mortality and migration), behavioral indicators (number and type of partners, condom use) and HIV program data (number of people on ART and PVT and the number of VMMCs conducted). The model is fit to data on prevalence from surveys, surveillance and routine testing by varying the epidemiological parameters within published ranges. [11] Ranges on the fitted values are used to generate uncertainty intervals on model output. The model is available for download free of charge from the Avenir Health web site as a module in the Spectrum software at https://avenirhealth.org/software-spectrum.php.

### Modeled populations

The Goals model calculates HIV incidence in the adult population between the ages of 15 and 49 using six categories for men (not yet sexually active, in a stable partnership, with multiple partners in the last year, clients of female sex workers, men who have sex with men and people who inject drugs) and five categories for women (not yet sexually active, in a stable partnership, with multiple partners in the last year, female sex workers and people who inject drugs). Each risk group is defined in terms of size and behaviors such as number of partners per year, acts per partner, condom use and needle sharing. Transition between groups is based on average duration within each group.

### Modeled HIV transmissions

Partners are chosen from within the same risk group except for those in stable partnerships where partners can be from any risk group depending on marriage rates. HIV transmission is determined by the number of partners, the number of contacts per partner, the probability of encountering an infected partner, and the probability of transmission per act adjusted for infected partner characteristics (stage of infection, type of sex, presence of another sexually transmitted infection and effective ART use) and susceptible partner characteristics (condom use, VMMC, presence of another sexually transmitted infection, clean needles and pre-exposure prophylaxis).

Incidence among adults aged 15-49 and age-specific incidence rate ratios are used to estimate incidence by age group (with strata spanning 15-19 and up to 80+) by fitting to age-specific prevalence from household surveys. New adult infections are tracked by age, sex, CD4 count and ART status. HIV-related mortality is determined by CD4 count, age, sex, and ART status. New child infections are determined from vertical transmission and HIV-infected children also are followed by CD4 category, sex, age, and ART status. New orphans are estimated from adult deaths and previous fertility and mortality trends. Full details of the model are provided elsewhere. [12]

### Historical analysis

We implemented the Goals model for 53 countries which account for 99.9% of PEPFAR spending. To estimate the historical impact of PEPFAR programs, we created a counterfactual scenario for each country by subtracting the PEPFAR services from the actual coverage of the five modeled interventions. Comparing the number of child new infections, child HIV-related deaths and AIDS orphans in the actual historical epidemic from 2004 to 2022 to the modeled estimates from the counterfactual analyses provides an estimate of PEPFAR’s historical impact. (For infections and HIV-related deaths, child is defined as <15 years of age. AIDS orphans include children ≤18 years.)

### Prospective analysis

For the prospective analysis we projected the epidemic from 2022 to 2030 under the assumption of constant coverage of all interventions at the 2022 level (reflecting a mix of PEPFAR and non-PEPFAR). To model the counterfactual (assuming immediate and complete PEPFAR cessation in 2024), we subtracted the 2022 PEPFAR services from the anticipated 2024 coverage in the absence of PEPFAR for the five modeled interventions and carried that coverage level forward through 2030.

## Results

### Historical analysis

The historical analysis indicates that from 2004-2022 the PEPFAR program averted 2.8 million new child HIV infections (Figure 1) and 1.5 million child HIV-related deaths (Figure 2). Without PEPFAR, by 2022 there would have been 8.2 million more AIDS orphans (Figure 3) and 1.0 million additional children living with HIV. The impact of PEPFAR is seen as soon as the program started but the impact becomes much larger as the effects have cumulated to 2022. The country-specific results are shown in Table 1.

**Figure 1.**
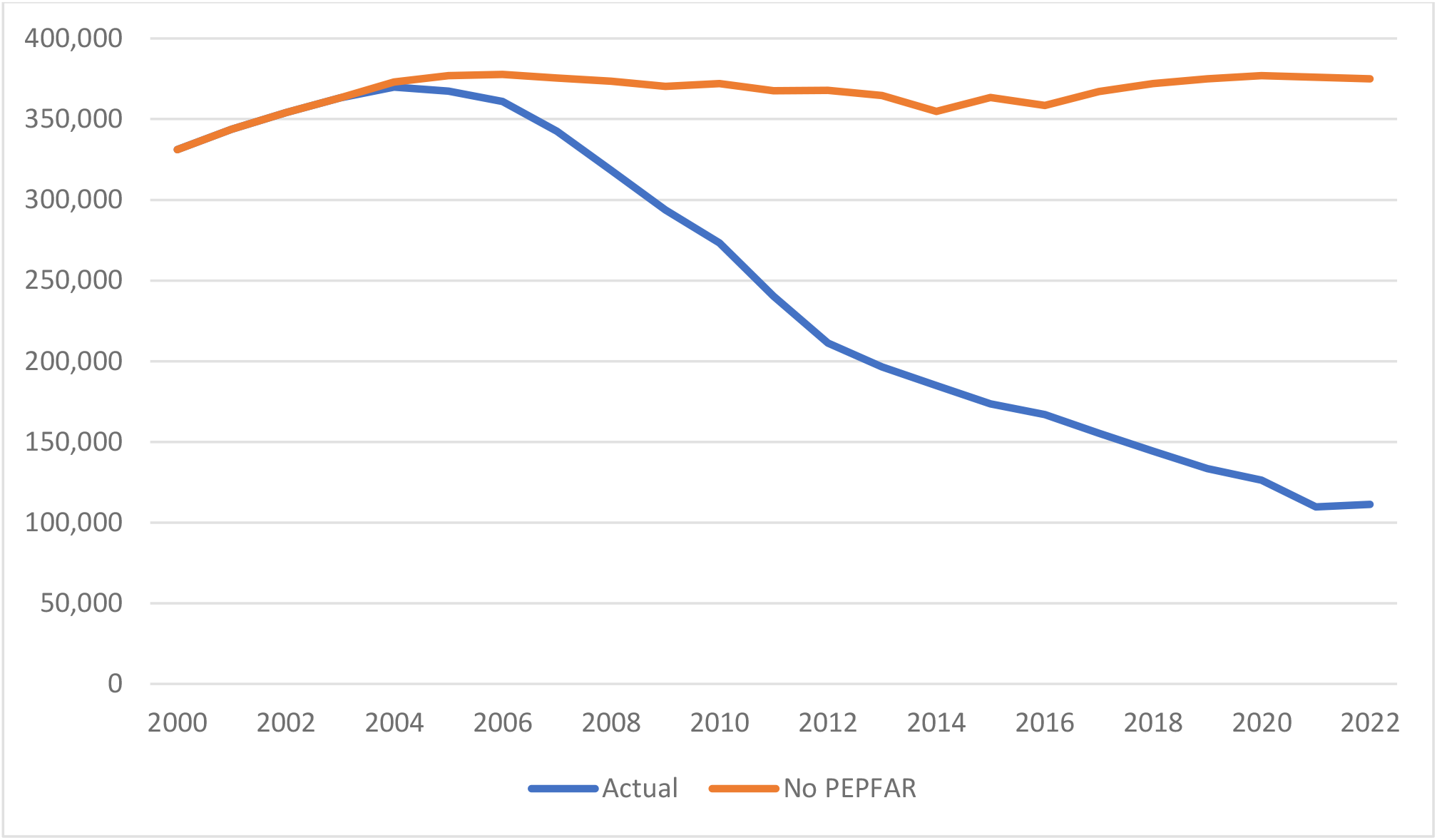
Number of new child HIV infections with and without PEPFAR, per year, 2000-2022

**Figure 2.**
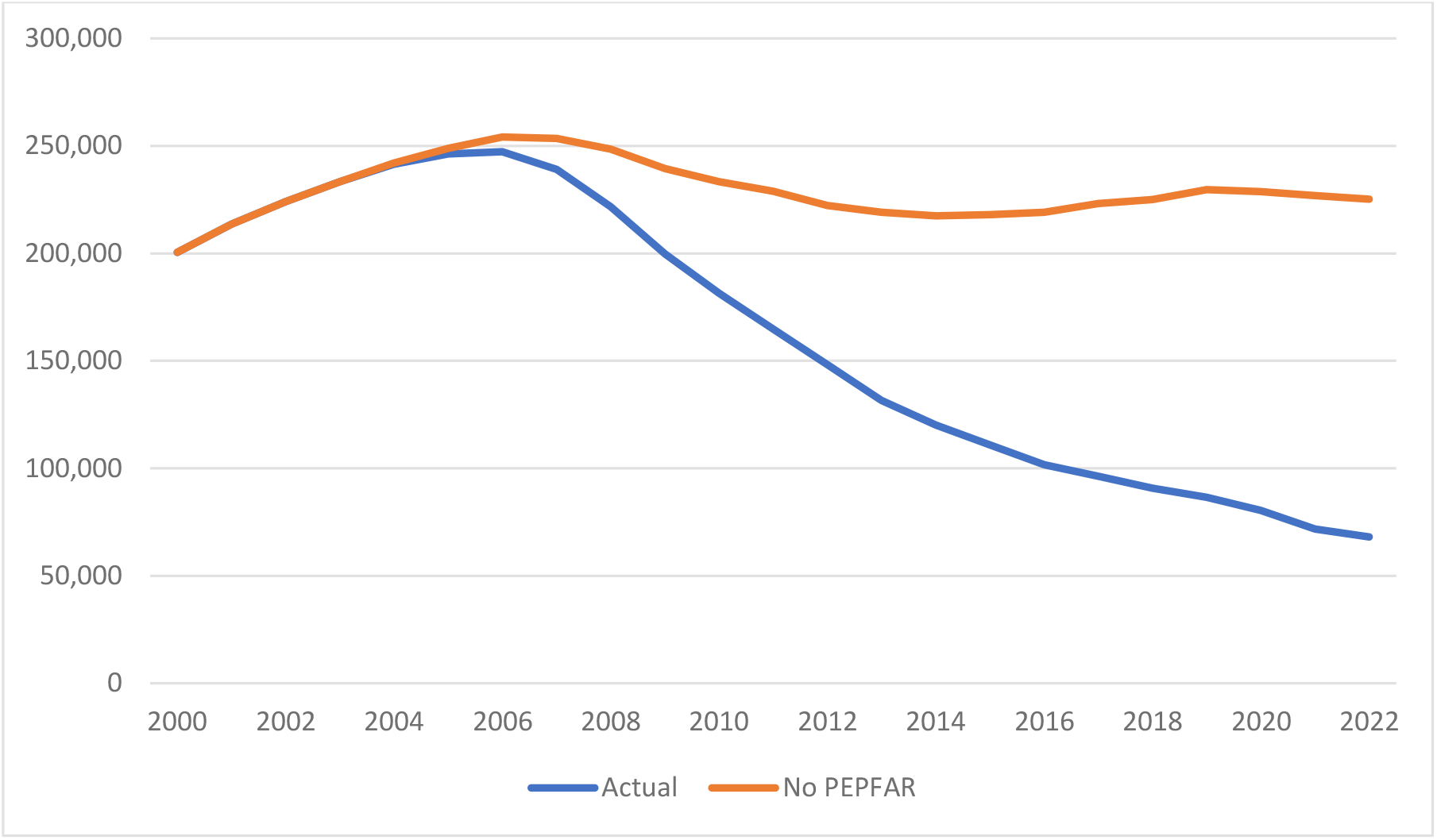
Number of child AIDS-related deaths with and without PEPFAR, per year, 2000-2022

**Figure 3.**
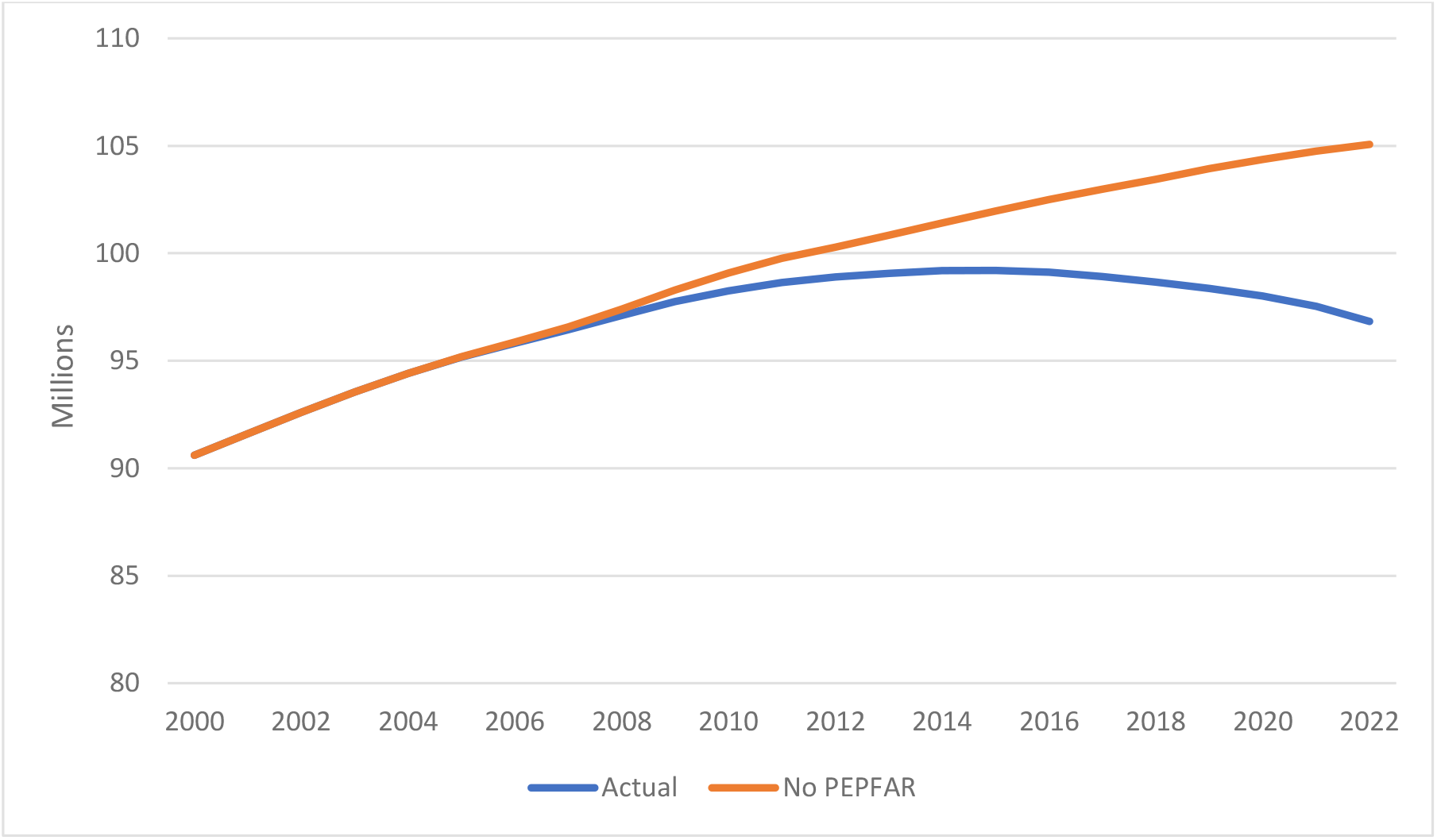
Number of AIDS orphans with and without PEPFAR, per year, 2000-2022

**Table 1.**
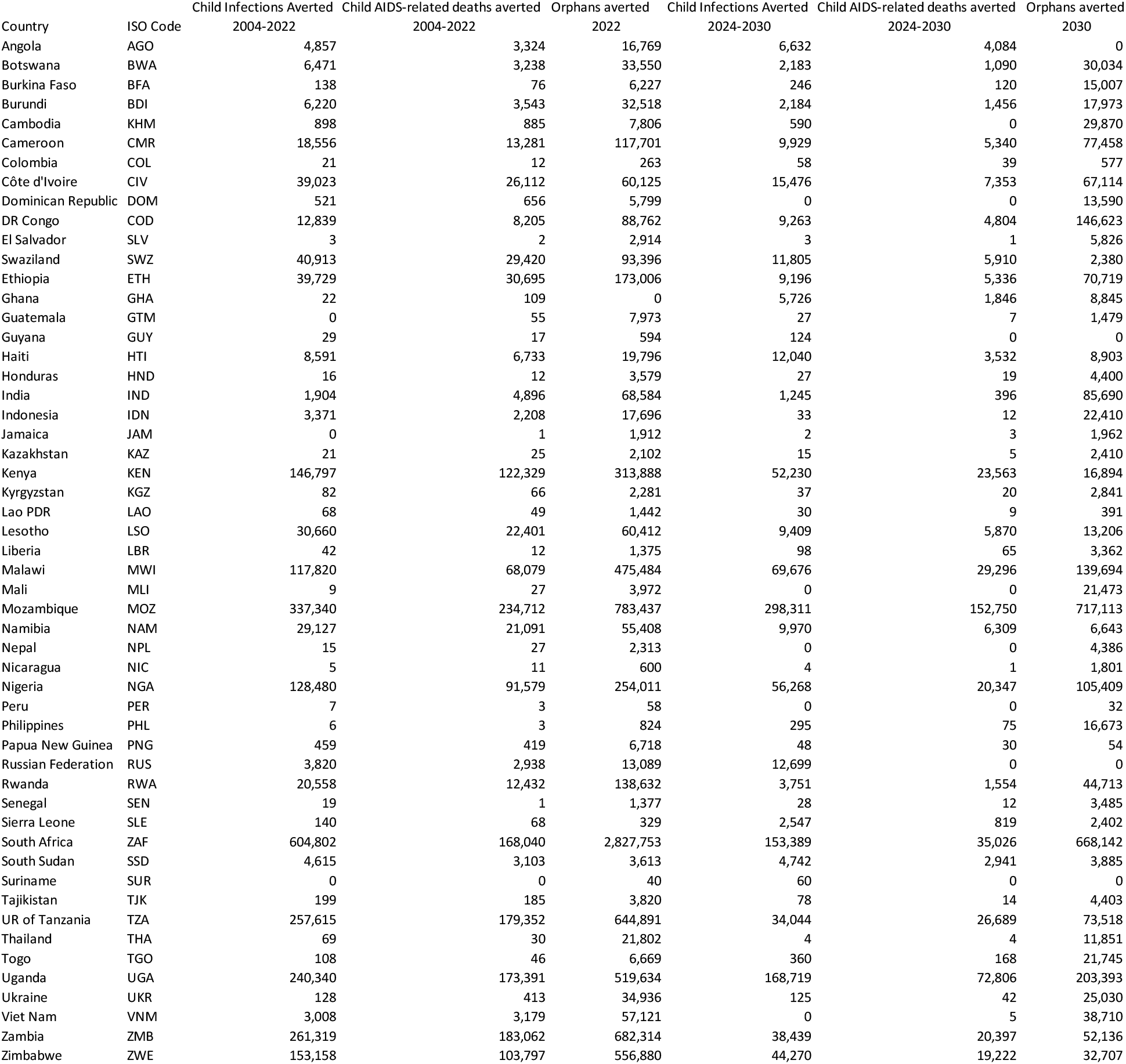
Results by Country.

### Prospective analysis

The prospective analysis indicates that a continuation of PEPFAR at its current level through 2030 would avert 1.03 million new child HIV infections and 460,000 child HIV-related deaths compared to a program cessation in 2024. In 2030 there would be 2.8 million more AIDS orphans without PEPFAR and 560,000 more children living with HIV. The continuation of PEPFAR at 2022 levels would facilitate service volumes from 2024-2030 of 7.3 million person-years of PVT, 200 million person-years of ART, 20 million VMMCs and 39 person-years of key population prevention services.

## Discussion

The PEPFAR program has been vital to the progress made to date in the global fight against AIDS. In addition to being the largest international donor, PEPFAR has provided leadership in scaling up interventions nationally, introducing innovations in service delivery and program monitoring and focusing efforts on the most cost-effective interventions delivered to the most at-risk populations and geographies. The impact of PEPFAR has been enormous. This contribution has also resulted in a current obligation. In 2022 PEPFAR supported 20 million people on ART and provided PVT services to 730,000 pregnant women. An immediate cessation of PEPFAR support would result in as many as 20 million AIDS-related deaths worldwide. PEPFAR’s ART, PVT and orphan support programs are vital to the health of tens of millions of adults and children in low- and middle-income countries.

There are several limitations to this analysis. We have assumed that no other sources of funding would substitute for the loss of PEPFAR funding. This assumption may be reasonable since PEPFAR is by far the largest international donor and PEPFAR accounts for 80% or more of all AIDS funding in some high-burden countries. The Goals model is fit to the same data used for the official UNAIDS estimates (generated by the Spectrum/AIM model), but because the underlying structures are different estimates of new infections and AIDS deaths are not identical. The results presented here rely on estimates of intervention effectiveness which are derived from studies, research cohorts and patient data. [13] There are uncertainties in each of these sources that need to be recognized. Moreover, these results were generated with a single model. Other models could have been used and would have produced somewhat different results. However, studies by the HIV Modeling Consortium have shown that while these models differ in some aspects, in general, they produce very similar results for incidence and mortality trends, especially during the historical period. [14-15]

There are many other prospective scenarios that we might have examined such as a gradual reduction in PEPFAR funding or the elimination of some but not all PEPFAR-supported interventions. Thus, our results may represent maximum impact. However, we only estimated deaths averted through 2030. Many of the children newly acquiring HIV between 2024 and 2030 would die of HIV-related causes after 2030 without PEPFAR support.

PEPFAR supports much more than the five interventions examined here. Crucially, it also provides support services for over 7 million orphans, a particularly vulnerable group that is characterized by lower life expectancy and higher rates of HIV acquisition. [16] PEPFAR also supports vital data collection, research and health system strengthening, which is crucial in addressing health conditions far beyond HIV/AIDS.

US funding for HIV sets an example for other donors. Large cuts in PEPFAR funding could also affect funding from other sources, especially through the Global Fund, resulting in even greater negative impact.

Through PEPFAR the United States has set an example of what can be achieved by a well-resourced and highly efficient public health program. PEPFAR has been particularly impactful among pediatric populations, having saved many lives of children and their parents. PEPFAR is now integral to the continued well-being of millions of people. And while the immediately impacted population is concentrated within low- and middle-income countries, the COVID-19 pandemic has demonstrated only too well that infectious diseases do not respect geographic borders. In ending PEPFAR, the United States would be abandoning its commitment to the world’s most vulnerable populations. Not only that, the end of PEPFAR would cripple efforts to end the HIV epidemic, which could have far-reaching impact felt throughout our globally connected world.

## Data Availability

All data produced in the present study are available upon reasonable request to the authors

